# Reduced motor planning underlying inhibition of prepotent responses in children with ADHD

**DOI:** 10.1101/2022.04.29.22274325

**Authors:** Irene Valori, Letizia Della Longa, Alessia Angeli, Gustavo Marfia, Teresa Farroni

## Abstract

To flexibly regulate their behavior, children’s ability to inhibit prepotent responses arises from cognitive and motor mechanisms that have an intertwined developmental trajectory. Subtle differences in planning and control can contribute to impulsive behaviors, which are common in Attention Deficit and Hyperactivity Disorder (ADHD) and difficult to be assessed and trained. We adapted a Go/No-Go task and employed a portable, low-cost kinematic sensor to explore the different strategies used by children with ADHD or typical development to provide a prepotent response (*dominant* condition) or inhibit the prepotent and select an alternative one (*non-dominant* condition). Although no group difference emerged on accuracy levels, the kinematic analysis of correct responses revealed that, unlike neurotypical children, those with ADHD did not show increased motor planning in non-dominant compared to dominant trials. Despite motor control could have compensated and led to good accuracy in our simple task, this strategy might make inhibition harder in more naturalistic situations that involve complex actions. Combining cognitive and kinematic measures is a potential innovative method for assessment and intervention of subtle differences in executive processes such as inhibition, going deeper than is possible based on behavioral outcomes alone.

**Significance Statement:** This study proposes an innovative method that integrates kinematic measurement with neuropsychological evaluation, thus providing information on the planning and control mechanisms underlying behavioral outcomes. It is applicable to the study not only of inhibition but more generally of executive functions, which are the basis of the ability of children and adults to achieve goal-directed actions. The use of a wearable motion sensor ensures good applicability in research, clinical evaluation, and intervention.

## Introduction

Performing cognitive operations and motor actions can be considered two faces of the same coin, as they vastly rely on shared mechanisms that allows us to produce appropriate responses with respect to goals and context (1). All relevant processes specialize with age, with motor and cognitive development being closely connected and inter-related in a dynamic process of exploring and adjusting to the to the demands of the external physical and social environment (2). Although cognitive and motor difficulties often co-occur in neurodevelopmental conditions and have been extensively studied as separate processes (3, 4), their common underlaying mechanisms are still to be furthered. We strongly believe that an integrate approach will provide a more complete understanding of the interplay between low-level sensorimotor processes and high-level executive functioning. Indeed, executive functions are those top-down processes (i.e., working memory, inhibition, shifting) that enable people to plan, monitor and control sensorimotor, socio-affective and cognitive processes, being fundamental to mental and physical wellbeing (5). Among these functions, the ability to inhibit automatic and highly probable responses, and let less probable alternatives successfully compete for control of cognition and behaviors, ensures that we are flexible and open to learning from the surrounding environment (6). Different paradigms are commonly used to measure the inhibition of prepotent responses (e.g., Stroop, Stop-signal and Go/No-Go tasks), with diverse versions that rely on mainly cognitive processes or entail varying degrees of motor components, and activate both distinct and shared neural areas (7, 8). For instance, cognitive inhibition of prepotent responses is conceived as the ability to focus on the task and ignore irrelevant distractors, as in the case of reading the world “blue” written in red ink. The motor component comes into play when the response requires some sort of movement (from pressing a button to reach a target), which sometimes has to be voluntarily stopped before or during its execution (7). Usually, these motor executions are not main targets of study, as they are considered only a way to obtain from individuals a response that is believed to reflect certain cognitive mechanisms. However, the very planning of this motor response could reveal important information about the processes at play. Thus, a deeper understanding of motor responses in cognitive tasks needs an improved consideration, leading to a new perspective on the shared mechanisms that underpin adaptive behaviors.

Inhibition of prepotent responses is a well-studied process being affected by disorders such as Attention Deficit and Hyperactivity Disorder (ADHD) (9), which is diagnosed based on inattentiveness, impulsiveness and hyperactivity symptoms (10). At the cognitive level, it is established that people with ADHD, despite the wide variability that characterizes developmental trajectories, are overall impaired in executive functions (11). At the motor level, it is still debated whether motor signs of atypical development can be detected from infancy and interpreted as early risk factors for the following development of ADHD cognitive and behavioral symptoms (12). Some co-occurrent difficulties in motor skills (e.g., fine motor precision, manual dexterity, bilateral coordination, balance, and postural control, running speed and agility, limb coordination, strength) can be found in about 50% of individuals with ADHD (13). However, those are not a diagnostic criterion and there is no evidence so far supporting the link between motor impairments and ADHD-specific symptoms such as inhibitory deficiencies (13). To shed light on this, an approach that studies these two aspects in an integrated manner could provide an innovative perspective on difficulties with inhibition and behavioral hyperactivity. Potential underlying mechanisms of inhibition difficulties relate to motor planning, which is responsible for selecting the action target and the timing of movements (e.g., reaction times, movement times, and acceleration/velocity parameters) (14). Adults with ADHD have been found to show atypical motor profiles, with longer reaction times to start moving after a “Go” cue and higher variability in the velocity shape along time, suggesting impaired motor planning capacities (15). It is interesting to note that there is a kind of slowness in sensorimotor and cognitive processes that underlie behavioral manifestations of impulsivity, hyperactivity, and inattention. A developmental perspective is needed to understand how these atypicalities have emerged and are maintained from childhood to adulthood. This would help us design targeted and age-appropriate interventions to promote a change on the mechanisms underlying the cognitive and behavioral difficulties of ADHD. Notably, purely cognitive training specifically targeting executive functions such as working memory, attention, inhibition, and shifting rarely result in cognitive nor behavioral or academic improvements, with scarce effect on ADHD core symptoms (16, 17). It has been speculated that leveraging embodied cognition and cognitive-motor approaches could boost training efficacy (18). This multidimensional perspective would eventually chart the way to define and test both motor and cognitive interventions to strengthen inhibition by passing through multidimensional doorways. Despite their presence and impact, motor difficulties of people with ADHD often end up being overlooked by research and clinical practice. Previous studies mainly based on correlational analysis of motor skills and purely cognitive performance at inhibition tasks, and failed to find clear relationships (13). On the other hand, investigating inhibition without dissociating motor and cognitive aspects that are deeply interrelated offers further insights on the underlying processes. The compelling possibility of integrating a kinematic measure to the traditional neuropsychological evaluation is strongly limited by the need of sophisticated motion capture systems. Those used for research purposes are often expensive and bulky, thus being hardly affordable for most clinical centers. In order to use low-cost portable solutions and boost the applicability of motion analysis, inertial sensors have been recently recommended for their good measurement reliability and validity (19). Adopting this technology in clinical practice would allow for a more detailed analysis of the mechanisms underlying the child’s performance on tests of interest. It could be used during assessment for setting specific intervention goals, for monitoring treatment effects, and as a treatment tool itself when used as biofeedback.

The present study aims at investigating children’s ability to inhibit prepotent motor responses, through an adapted version of the Go/No-Go paradigm. The task embedded a reaching movement and kinematic measures to surface motor planning characteristics of inhibition (20). Children with ADHD or typical development were recruited. A commercially available, low-cost, easy to use, wearable accelerometer sensor was employed to capture movement features. Distinct kinematic indices were considered to study the progress of motor planning in the various phases of action. Reaction Time (RT), from the appearance of the Go stimulus to the beginning of the movement, gives a measure of pure motor planning. Higher need for motor planning is expected to results in higher RTs (21). This is the index of choice for studying variability in the inhibitory abilities of people with ADHD (22). During movement execution (as measured by the Movement Duration - MD), motor planning gradually gives way to control and monitoring of the ongoing movement, which involve distinct processes (14). Therefore, the percent Time to Peak Velocity (TPV) may represent a useful index to disentangle how much of the movement time is devoted to planning or control. Theoretical (e.g., in robotics) reaching trajectories starting and ending at full rest will show a bell-shaped velocity path, with the first half of MD spent accelerating and the second one decelerating, resulting in a 50% TPV (23, 24). The more cognitive load is required in a given task, the more human reaching movements have a greater need for motor planning, thus resulting in increased acceleration phase and TPV (20).

We have previously found that, to correctly inhibit a prepotent response and select the instructed alternative one, neurotypical adults show longer RT and MD, as well as increased TPV overall dedicating more resources to motor planning than monitoring and control of ongoing movements(20). Assuming that this is the motor strategy that has been established as most effective in adults, a developmental perspective is needed to understand how it specializes during childhood and is potentially subject to deviation in cases of atypical development. We therefore expect age-related differences in the kinematic profile of motor planning and control necessary to inhibit prepotent responses. Moreover, we hypothesize that children with ADHD, compared to neurotypical controls, would show greater difficulties inhibiting the prepotent response, which the literature also refers to as motor impulsivity (25). We expect children with ADHD to make more errors than controls in the non-dominant condition, and show an atypical motor profile, with reduced or less effective motor planning. As markers of motor impulsivity, we particularly expect reduced RT and TPV in the group of children with ADHD (25).

## Results

To analyze children’s performance, we considered 4 dependent variables. Accuracy indicates the percentage of correct responses out of the total number of valid responses (after discarding anticipations and omissions). RT measures the time from the appearance of the central stimulus to the onset of movement (the time when the hand is raised by the presence sensor). MD measures the duration of the movement (from when the sensor is released to when a response is given). We then computed the percent Time to Peak Velocity (TPV), which is the percentage of MD spent from movement onset to maximum peak velocity. Data calibration and pre-processing of raw acceleration data, as well as computation of TPV values were conducted following the steps reported in our previous work (20). Participants provided 6,576 valid responses. We excluded those responses whereby either RT or MD were less than 100 ms, being them ascribable to anticipation. We therefore included responses whereby the TPV was within the 5-95% range, thus considering extremes as due to extra-task movements. Filtered responses (217/6,576 = 3.3%) were removed and not further analyzed. Final dataset comprehended 6,359 observations.

An exploratory approach was elected to test different potential hypotheses linking each dependent variable to the predictors of interest. Through separated sets of model comparisons, different research hypotheses were specified as statistical models, and their statistical evidence was evaluated using information criteria (26). Generalized mixed-effects models were employed to account for the repeated measures design of the experiment (i.e., trials nested within participants), and specify the distribution of each dependent variable. For each dependent variable, a set of models were compared through the Akaike weights (AICcWt) (i.e., the probability of each model, given the data and the set of considered models) (26), using the *AICcmodavg* (26) R package. Then, likelihood ratio tests were used to compare the chosen models, and test the effects predicted by the best model. These analyses have been run in R (27).

We separately investigated whether each dependent variable (Accuracy, RT, MD, TPV) was influenced by the fixed effects of Condition (within-subjects, two levels categorical factor: dominant versus non-dominant), Group (between-subjects, two levels categorical factor: ADHD versus TD), and Age (continuous numeric variable). All models accounted for the random effect of participants (i.e., interpersonal variability). We considered the five models that follow.

- **m0** (null model) specified the hypothesis of no difference due to the independent variables and only accounted for individual variability
- **m1** specified the hypothesis of a Condition effect
- **m2** specified the hypothesis of additive Condition and Group effects
- **m3** specified the hypothesis of additive Condition, Group and Age effects
- **m4** specified the hypothesis of a two-way interaction effect between Condition and Group, with the additive Age effect.

### Accuracy

Children with ADHD provided 2,234 correct and 137 incorrect responses. TD children provided 3,777 correct and 211 incorrect responses (percentages of correct responses according to Group and Condition are reported in Table S1, SI Appendix). Model comparison was run with the *glmmTMB* (28) R package. The binomial distribution was specified to account for the binary nature of the dependent variable (1 = correct; 0 = incorrect). According to AIC, the best model was **m1** (AICcWt = 0.39; χ^2^ = 369.3; p < .001), which revealed a significant effect of Condition (p < .001). As visualized in Figure 1, accuracy was reduced in the non-dominant condition.

**Figure 1.**
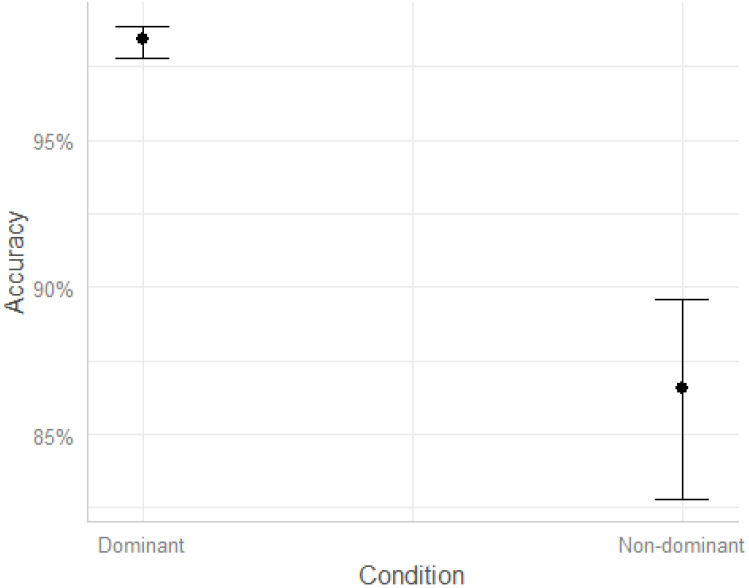
Predicted effect of Condition on Accuracy (n_trials_ = 6,359, n_ADHD_ = 17, n_TD_ = 26).

### Kinematics

We further explored kinematic features of correct responses to investigate whether, beyond accuracy, children with ADHD would show subtle motor atypicalities. Means and standard deviations of RT, MD, and TPV of correct responses in each condition and group are reported in Table S2, SI Appendix.

#### RT

Model comparison was run with the *glmer* function of *lme4* (29) R package. The gamma distribution was specified to account for the positively skewed nature of the dependent variable. According to AIC, the best model was **m4** (AICcWt = 0.80 χ^2^ = 4.9; p = .03), which revealed a significant interaction between Condition and Group (p = .03), and a significant effect of Age (p < .001). As visualized in Figure 2, TD children showed increased RT in the non-dominant compared to the dominant condition, thus devoting more time to motor planning when the response required inhibition. This pattern was not present in children with ADHD, who did not differentiate RT depending on Condition. Moreover, there is a negative association between RT and Age, with RT decreasing at older ages, regardless of group.

**Figure 2.**
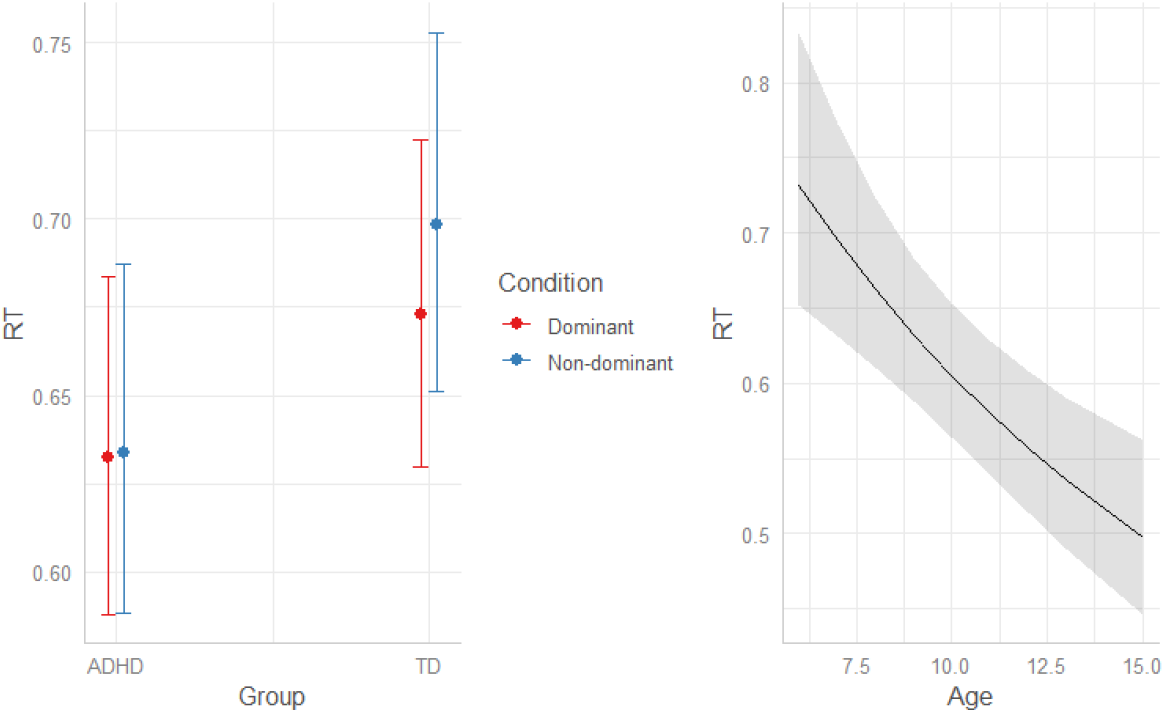
Predicted effects of Condition*Group and Age on RT (n_trials_ = 6011, n_ADHD_ = 17, n_TD_ = 26).

#### MD

Model comparison was run with the *glmer* function of *lme4* (29) R package. The gamma distribution was specified to account for the positively skewed nature of the dependent variable. According to AIC, the best model was **m2** (AICcWt = 0.41 χ^2^ = 2.7; p = .1), which revealed a significant effect of Condition (p < .001), and a non-significant effect of Group (p = .09). As visualized in Figure 3, MD increased in the non-dominant condition and tended to be reduced in TD compared to ADHD children.

**Figure 3.**
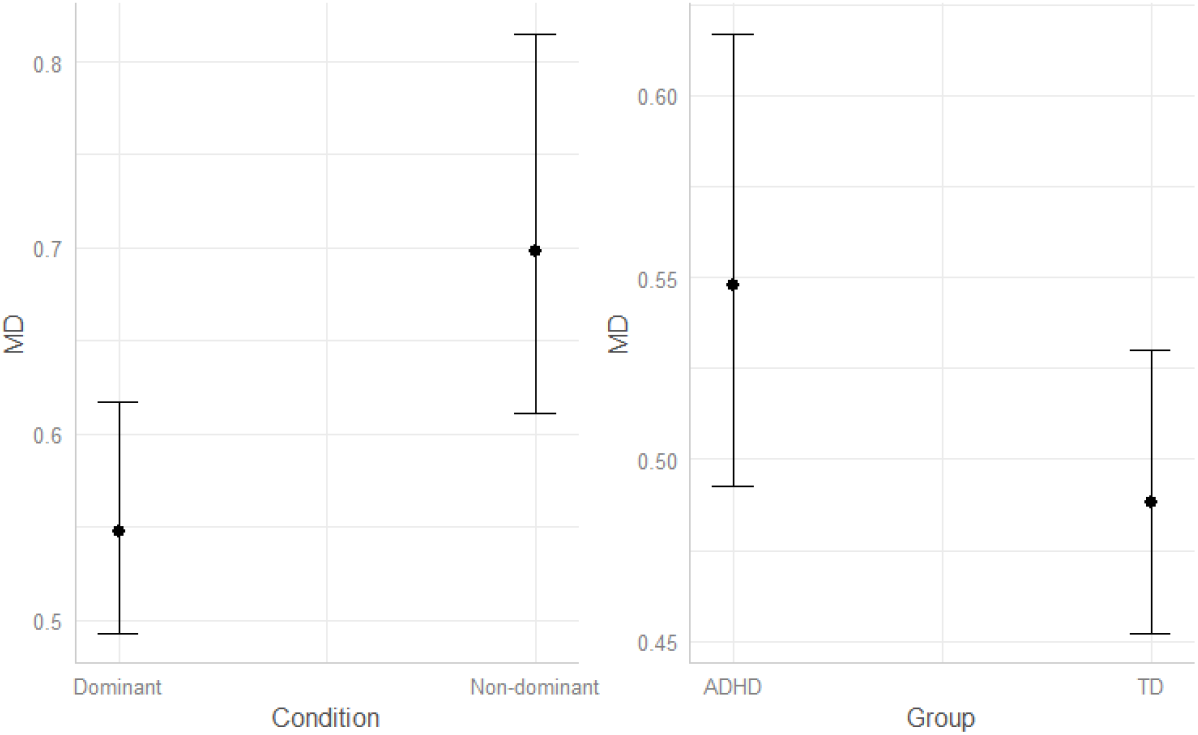
Predicted effects of Condition and Group on MD (n_trials_ = 6011, n_ADHD_ = 17, n_TD_ = 26).

#### TPV

Model comparison was run with the *glmmTMB* (28) R package. The beta distribution was specified to account for the nature of the dependent variable (continuous proportions on the interval 0:1). According to AIC, the best model was **m4** (AICcWt = 0.83 χ^2^ = 8.3; p = .004), which revealed a significant interaction between Condition and Group (p = .004), and a non-significant effect of Age (p = .3). As visualized in Figure 4, TD children showed increased TPV in the non-dominant compared to the dominant condition, thus devoting more time to motor planning when the response required inhibition. This pattern was not present in children with ADHD, who did not differentiate TPV depending on Condition. At both the group and individual level, further graphical inspection of velocity shape across time is described in SI Appendix.

**Figure 4.**
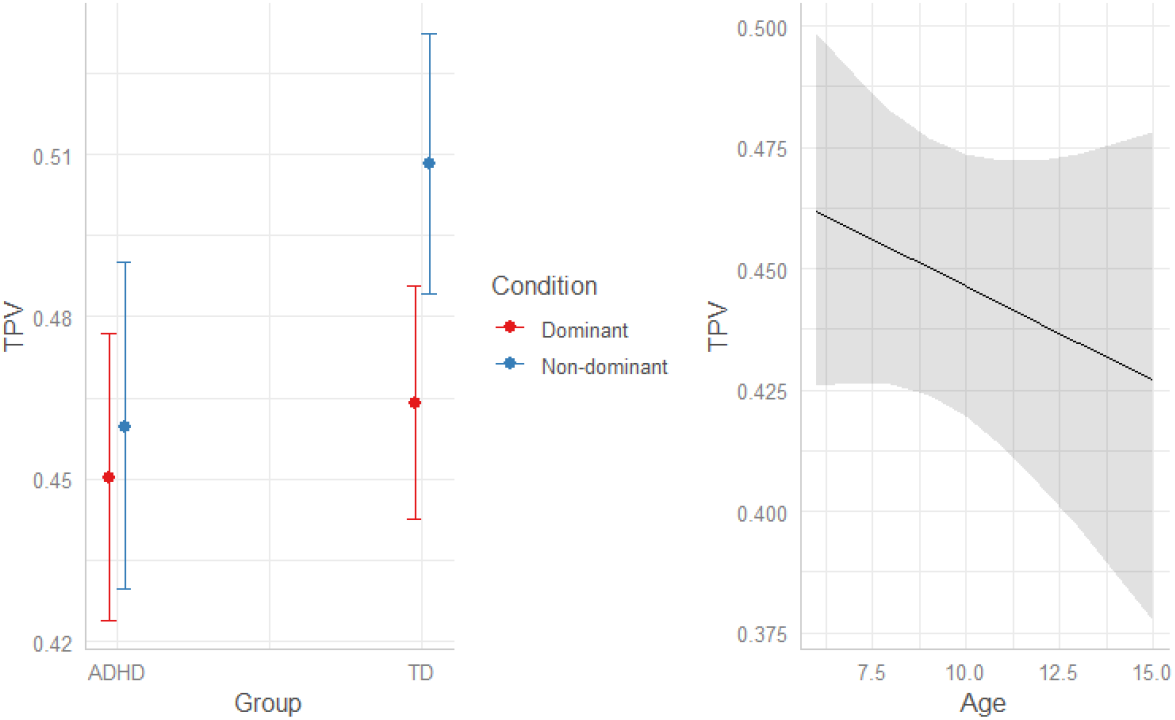
Predicted effects of Condition*Group and Age on TPV (n_trials_ = 6011, n_ADHD_ = 17, n_TD_ = 26).

## Discussion

The present study explored the mechanisms underlying the inhibition of a prepotent motor response, which is frequently reported to be affected in children with ADHD. The performance of the ADHD and TD groups at our motor adaptation of the Go/No-Go task, showed both similarities and differences.

Both ADHD and TD children made more errors in the non-dominant compared to the dominant condition. This indicates that the task was effective in inducing a prepotent response in the dominant condition, which was the more frequent one, and facilitated by the requirement to match the Go stimulus and the response option by color. Children with ADHD and typical development were equally accurate in selecting the correct response, so that no group difference was found on accuracy levels. This unexpected result could be due to the ease of the task, which required a rather simple motor response, as also evidenced by the high percentages of correct responses (i.e., potential ceiling effect). In tasks with greater time pressure or greater complexity of the motor action required to answer, we could expect more marked differences between the two groups. We can also see a progressive reduction in RT as the age of the participants increases, which is consistent with decades of findings from developmental studies (30). This suggests that motor planning becomes globally more effective and rapid with age, and therefore requires fewer cognitive resources.

The main findings of this study revealed that, beyond accuracy, the ADHD group showed different motor patterns that possibly indicate reduced motor planning compared to the TD group. In the non-dominant condition compared to the dominant condition, TD children spent more time planning the movement, which resulted in longer Reaction Time (RT) and greater percent Time to Peak Velocity (TPV). On the other hand, children with ADHD did not modulate these two indices according to condition, not dedicating more time to motor planning when needed to inhibit the prepotent response. This subtle lack of flexibility in adjusting the motor and cognitive strategies to the task demands can be interpreted as a marker of motor and cognitive impulsivity. Despite being non statistically significant, additional tendencies of our kinematic data are worth being described. We noticed that children with ADHD, compared to controls, tended to have longer MD, potentially indicating that they are controlling their movement along the way, instead of preplanning. This is also suggested by the smaller TPV captured in the ADHD group across conditions, with higher portion of movement being dedicated to the deceleration phase, that usually stands for motor control (14). We might speculate that children with ADHD employ different motor and cognitive strategies, with greater reliance on ongoing monitoring and readjustment than planning of movements and actions. This result can contribute shedding light on previous findings that reported increased movement variability in children with ADHD (31). This has often been interpreted as an indication of poor motor control, when instead it could be a compensatory strategy that, given a reduced planning, requires more online adjustments during movement execution. Online control might help children with ADHD compensate for planning difficulties, which may be sufficient to achieve good accuracy in very simple tasks as the one employed in our work. Indeed, they chose between two alternatives that differed only in one motor (i.e., the movement direction: reaching the key to the right or to the left of the central stimulus) or cognitive (i.e., the response key color) parameter. However, this might not be sufficient in more naturalistic situations, in which alternative choices differ in more complex kinematic parameters (e.g., using the right arm or the left arm to respond), or require finer cognitive processing (e.g., selecting the most appropriate behavior according to a specific social context).

In everyday life, children constantly perform actions that require planning and control, as well as inhibition of automatic behaviors as the demands of their environment change. Further research is needed to investigate the implications of atypical motor and cognitive inhibition on the daily life, learning, and social skills of children with ADHD. For instance, some of the children in our ADHD sample were reported to show stereotypies, which are involuntary, restricted and repetitive patterns of behaviors that limit the child’s resources to learn and practice various, appropriate and goal-directed actions (32–34). Specifically, motor stereotypies are present in both neurodevelopmental conditions and typical development (35), and might be related to ineffective motor planning (36) and inhibitory difficulties (37). Indeed, motor-related cortical potentials in premotor areas, which anticipate voluntary motor actions, are found to be absent before stereotypy onset in typical development (36). Stereotypies are mostly studied in Autism Spectrum Disorders (ASD), as they are core symptoms of those conditions (10). However, they are frequently found in ADHD, and show similar characteristics across ASD and ADHD (38), which often co-occur, share clinical manifestations, and entail impairments in overlapping mechanisms (39, 40). In addition, studies on ASD suggested that atypical inhibition of prepotent responses is correlated with repetitive behaviors, with differences between higher-order (preoccupations, restricted interests, compulsive routines, ritualistic behaviors) and sensorimotor (repetitive movements and sensory preoccupations) stereotypies (41, 42). Moreover, stereotypies are associated with sensory difficulties in children with ASD (43). Remarkably, children with ADHD may also present atypical sensory processing (44–46), which is bounded to motor and cognitive processes through complex, dynamic, and multidirectional relationships. We can speculate that those children with greater stereotypies and less effective sensory and executive profiles could have reduced motor planning abilities, and therefore need to devote more resources to motor control to effectively inhibit a prepotent response. Future studies may employ our paradigm to better understand whether atypical cognitive and motor inhibition may contribute to broader individual differences in everyday sensory, cognitive, and social functioning. Studies with more hypothesis-driven approaches and appropriate sample size would allow to draw clearer, more inferential conclusions on the complex relationships between these variables.

This study opens the door to important application challenges in bringing these methods and knowledge into clinical practice. It would be crucial to integrate the kinematic analysis to the classical neuropsychological tests that evaluate executive functions, to better understand how a response to a given test is planned and adjusted along the way. This method would facilitate not only the identification of specific difficulties and the monitoring of the treatment effects, but also serve as an intervention tool itself. For instance, using kinematic measures as biofeedback could promote patients’ awareness of their behaviors and facilitate learning strategies to modify them. Although the use of inexpensive and portable kinematic sensors removes one of the barriers to its use in the clinic, the difficulty of analyzing and interpreting the raw data obtained with such instruments remains. To overcome this obstacle, it will be necessary for researchers to develop and make available user-friendly software that process the raw kinematic data and calculate performance indices that are interpretable by clinicians. To this end, we first need large-scale validation studies that provide normative values and risk indices to evaluate an individual’s performance.

It is worth mentioning that the present study has some limitations. As we were not interested in assessing gender differences, our sample is not balanced by participant gender, which reduces its representativeness of the general population. In addition, the sample size was determined by the number of families that agreed to participate in the study. Given the complexity of the experimental design (i.e., multiple dependent and independent measures are of interest), its exploratory nature, and the paucity of prior evidence on which to estimate expected effect sizes and appropriate sample sizes, our sample size may be insufficient to reveal further differences between groups. Further inferential research will be needed to confirm the considerations presented in this paper.

In conclusion, children with ADHD can exhibit similar accuracy than neurotypical controls in simple tasks tapping on the inhibition of prepotent motor responses. However, accurate inhibition appears to be achieved through different mechanisms, including less motor planning and greater ongoing control of movements. Although online control of one’s own responses may be sufficient to compensate for planning difficulties in simple experimental tasks, this could profoundly impact the behaviour of children with ADHD in everyday life contexts, which involve very complex choices among numerous possible alternatives. Moreover, motor, and cognitive impulsivity might be related to broader atypicalities, ranging from sensory atypia and stereotypies to executive difficulties in everyday tasks. For this reason, it is fundamental to understand the mechanisms underlying impulsivity, design interventions that are individualized on the child’s profile and synergistically target the motor and cognitive dimensions of inhibition. To this end, the use of portable, user-friendly, and low-cost kinematic sensors offers great possibilities for neuropsychological assessment and treatment, being also affordable for local clinical services. In sum, this study opens the door to further research that will help the scientific and clinical community understand and target impulsivity, leading to benefits on children’s developmental trajectory and quality of life.

## Materials and Methods

### Participants

We recruited 17 children with ADHD (4 female children) from 6 to 15 years of age (M =9.4, SD = 2.2), and 26 children with Typical Development (TD control group; 10 female children), from 6 to 13 years of age (M = 9.2, SD = 2.1). Three additional participants (2 in the ADHD and 1 in the TD group) were excluded due to technical issues that prevented them from completing at least 50% of the trials.

Children with ADHD were recruited and tested at a clinical center located in the north of Italy. Psychologists confirmed children’s diagnosis and provided IQ assessments through the WISC-IV scale. Moreover, we collected parent-reported questionnaires on the child’s executive (Executive Functions Questionnaire – Q.FE (47)) and sensory profile (Short Sensory Profile – SSP (48)), as well as the presence and severity of restricted and repetitive behaviors (Repetitive Behavior Scale-Revised – RBS-R) (49). A convenient control group of children with typical development in the same age range was tested at the University of Padova. According to parents’ reports, typically developing children had no medical or neuropsychological conditions.

Characteristics of the ADHD group are provided in Table S3 of SI Appendix, which includes IQs, and scores from the parent reported assessment. Comorbidities are also described in SI Appendix. All children’s parents signed a written consent form. All experimental methods received ethical approval from the Research Ethics Committee of the School of Psychology, University of Padova (protocol no. 3251). The experiment was carried out in accordance with the approved guidelines and regulations.

### Procedure and task

Children sat on a desk and wore an accelerometer sensor on their dominant wrist. They were instructed to place the dominant hand at a specific starting position, monitored by a presence sensor, and completely extend their arm to tap on the response touchscreen. A Go/No-Go paradigm was adapted to assess the inhibition of a prepotent response and tested with neurotypical adults in a previous work(20). Upon comparison of a central stimulus (red/green, upwards/downwards arrow), participants were asked to select, reach, and press one of two response keys (either a red or green circle) placed one on the left and one on the right side of the central stimulus, following specific instructions. Before the start of the next trial, participants had to return their hand on the sensor. As soon as the hand was in place, the next trial started after a random delay (range = 0:2 seconds), which prevented children from anticipating the onset of the next trial. The set-up and procedure are illustrated in Figure 5.

**Figure 5.**
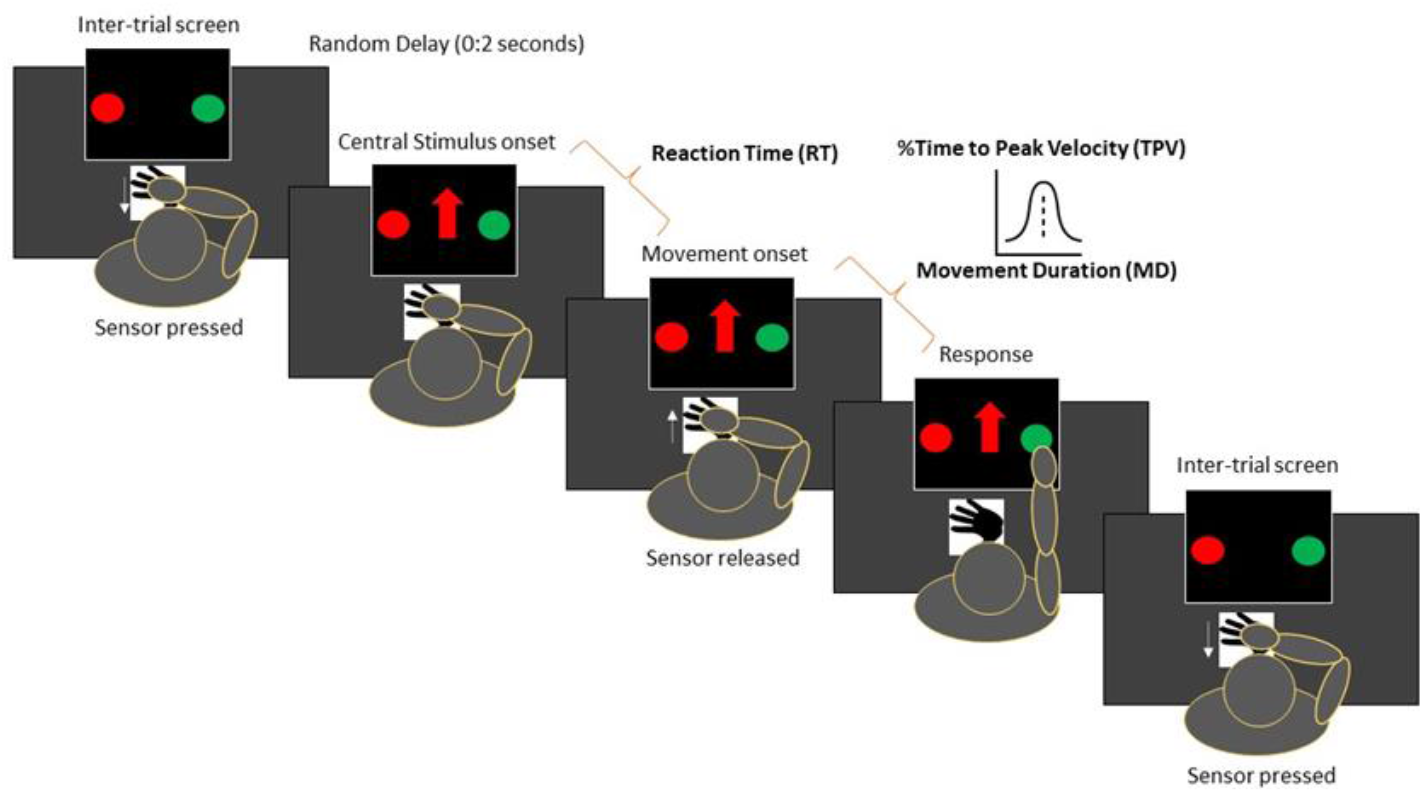
Set-up and procedure.

More in detail, participants were told to select the response key of the same color of the central stimulus when it was an upwards/downwards (counterbalanced between participants) arrow (*dominant condition*). On the other side, they were told to select the response key of the different color when the central stimulus was an averted (either upwards or downwards, counterbalanced between participants) arrow (*non-dominant condition*). We elicited a prepotent response for the same-color action (occurring the 75% of times), and an inhibitory response for the alternative different-color action (occurring the 25% of times).

Participants were instructed to reply as quickly and accurately as possible. Failure to press any keys within 2,000 ms was marked as “omission”. Movements starting before the cue stimulus onset were tagged as “anticipation” (the program aborted the trial by showing no cue stimulus). The task ended upon completion of 160 valid trials (i.e., trial with correct/incorrect answer) or 180 total trials. Two blocks were administered, with the red/green response keys being located once on the right and once on the left side of the touchscreen. To maintain participants’ engagement during the task, a short (30 seconds on average) video from well-known movies appeared every 40 trials. The task lasted about 15 minutes. Technical features of the apparatus (e.g., programming language, devices for conducting the experiment) are described in detail in our previous work (20).

## Data Availability

All data produced are available online upon publication at https://osf.io/gjha4/?view_only=6fa8fe8e2ee2442fb2df64e1dad06ac3

## Acknowledgments

We sincerely thank the psychologists Alessandra Cagnin and Donatella Benetti for their enthusiastic collaboration on this project. Our gratitude to the students Irene Guglielminetti, Krizia Indri, and Giulia Mantovani, who contributed to data collection. Many thanks to Andrea Janna for his technical support.

## Notes

### Competing Interest Statement

The authors have declared no competing interest.

### Funding Statement

This study did not receive any funding

### Author Declarations

The Research Ethics Committee of the School of Psychology, University of Padova gave ethical approval for this work (protocol no. 3251).

