## Supplementary material for "Reduced motor planning underlying inhibition of prepotent responses in children with ADHD": SI Appendix

### Supplementary Information for
Reduced motor planning underlying inhibition of prepotent responses in children with ADHD.

Irene Valori[1], Letizia Della Longa[1], Alessia Angeli[2], Gustavo Marfia[3] and Teresa Farroni[1,4*]

[1]University of Padova, Department of Developmental Psychology and socialisation, Padova, 35131, Italy
[2]University of Bologna, Department of Computer Science and Engineering, Bologna, 40126, Italy
[3]University of Bologna, Department of the Arts, Rimini, 47921, Italy
[4]Padua Neuroscience Center (PNC), University of Padova, Italy

Teresa Farroni


**This PDF file includes:**

Supplementary text
Supplementary Figures
Tables S1 to S3
SI References

#### Descriptive statistics

##### Accuracy

Children with ADHD provided 2,234 correct and 137 incorrect responses. TD children provided 3,777 correct and 211 incorrect responses (percentages of correct responses according to Group and Condition are reported in Table S1).

| | | Accuracy | |
|---|---|---|---|
| **Group** | **Condition** | Mean | SD |
| ADHD | Dominant | 98 | 3 |
| | Non-dominant | 82 | 16 |
| TD | Dominant | 98 | 2 |
| | Non-dominant | 85 | 11 |

**Table S1.** Descriptive statistics of accuracy levels, % of correct responses ($n_{ADHD}$ = 17, $n_{TD}$ = 26).

##### Kinematics

Means and standard deviations of RT, MD, and TPV of correct responses in each condition and group are reported in Table S2.

| | | RT | | MD | | TPV | |
|---|---|---|---|---|---|---|---|
| **Group** | **Condition** | Mean | SD | Mean | SD | Mean | SD |
| ADHD | Dominant | 652 | 211 | 565 | 217 | 447 | 182 |
| | Non-dominant | 653 | 217 | 734 | 242 | 456 | 228 |
| TD | Dominant | 691 | 198 | 514 | 190 | 460 | 175 |
| | Non-dominant | 716 | 217 | 656 | 242 | 504 | 215 |

**Table S2.** Descriptive statistics of correct responses, values in ms ($n_{trials}$ = 6,011, $n_{ADHD}$ = 17, $n_{TD}$ = 26).

#### Demographic information

Characteristics of the Attention Deficit and Hyperactivity Disorder (ADHD) group are provided in Table S3, which includes IQs, and scores from the parent reported assessment.

| | IQ | RBS Tot | Low-level RRB | High-level RRB | Q-FE Tot | SSP Tot |
|---|---|---|---|---|---|---|
| M | 107.2 | 21.8 | 7.8 | 14.0 | 90.6 | 136.4 |
| SD | 17.6 | 15.5 | 6.5 | 11.8 | 20.9 | 26.2 |

**Table S3.** ADHD group characterisation ($n_{ADHD}$ = 17). Mean (M) and Standard Deviation (SD) of:

IQ: total score from the WISC-IV scale. RBS Tot: total score from the RBS-R; higher scores indicate a more severe profile of restricted and repetitive behaviours. Low-level RRB: scores from Stereotyped, and Self-Injurious subscales of the RBS-R. High-level RRB: scores from Compulsive, Ritualistic, Sameness and Restricted Interests Behaviours subscales of the RBS-R. SSP Tot: total score from the SSP; higher scores indicate better sensory profile. Q.FE Tot: total score from Q.FE; higher scores indicate better executive functions.

Six children received a comorbid diagnosis of Specific reading disorders (from moderate to severe). Two children received a diagnosis of Specific spelling disorder (moderate), and four were diagnosed with other behavioural and emotional disorders.

### Velocity shape and trend

We here conduct a visual inspection of our data, with a specific focus on velocity shape and trend across movement time. Notably, only correct trials (i.e., trials in which the participant gave the correct answer) are considered. Firstly, we plotted the data of the two groups separately (**TD children** and **ADHD children**), then we plotted individual data to explore individual variability. As explained in our previous work [1], we do not look at velocity magnitude values, but rather focus on its curve shape and trend in time.

At the group level, each Figure is composed by 3 graphs, one for each row. The first and second graphs constitute of a boxplot composition from trials in either the dominant (red) or non-dominant (blue) condition, respectively. The x-axis represents the movement time (in ms), whereby the instant in which the participant starts moving is aligned with the 0 value. The y-axis shows the velocity values (in m/s). For each 10 ms of movement time (corresponding to the accelerometer sampling rate), we plotted a boxplot composed by data from all equivalent time points of the different trials. Although, the y-axis value ranges were affected by outliers, they were excluded from visualization for the sake of graphic clarity and readability. For instance, in case some blank spaces appear in the superior and inferior parts of a boxplot, some invisible outliers are present. As we focus on the velocity shape and trend across time, and do not aim to compare its magnitude across different graphs, we did not set a fixed y-axis range for all the Figures. We have therefore avoided a flattening of the boxplots resulting from variability between participants. As not all the trials have the same movement duration, the boxplots are composed by varying amount of data. We take this into consideration in the third graph, which represents the number of trials contributing to each time instant of each boxplot.

At the individual level, we also reported a plot representing velocity in all single trials. Each curve corresponds to a single trial and is visualized in red for the dominant condition, and in blue for the non-dominant condition. The x-axis represents the movement time (in ms), while the y-axis shows the velocity values (in m/s). In addition, a vertical green line marks the time instants when the participant ended its movement in each trial (namely, touched the response screen and provided an answer). Green lines make it easier to capture movement duration in each trial.

#### 1. TD and ADHD groups

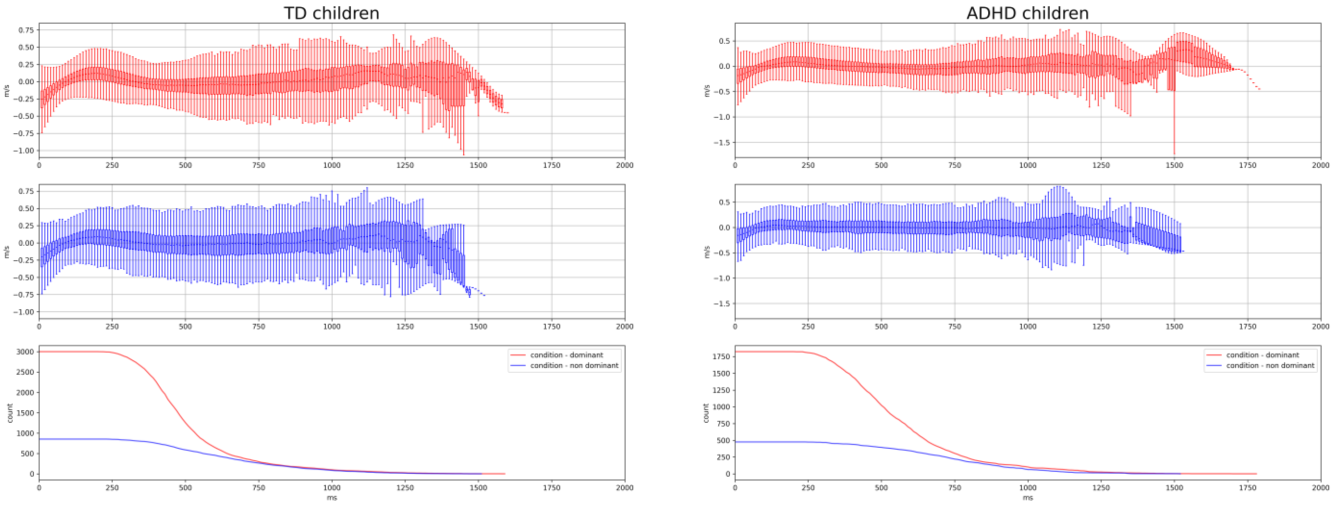


From a group level visualisation of the boxplot composition graphs, we observe that the **TD group** showed a more pronounced bell-shaped velocity pattern at the beginning of movements, as represented by the time-ordered set of boxplots. This seems in line with previous literature suggesting that children with ADHD do not show a typical bell-shaped velocity profile, which indicates impaired motor planning [2].

#### 2. TD children

Here we report graphical visualisations of individual data from the **TD group**. For this cohort of 26 participants, numbers ranging from 101 to 126 are reported in the Figure title as participants' identification code.

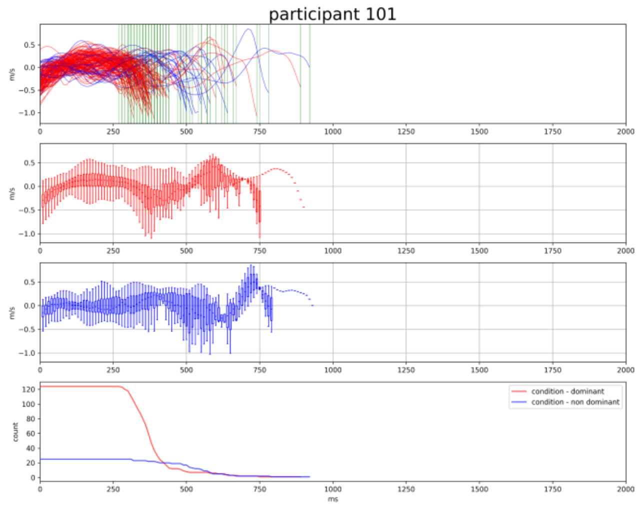
participant 101
m/s
count
ms
condition - dominant
condition - non dominant

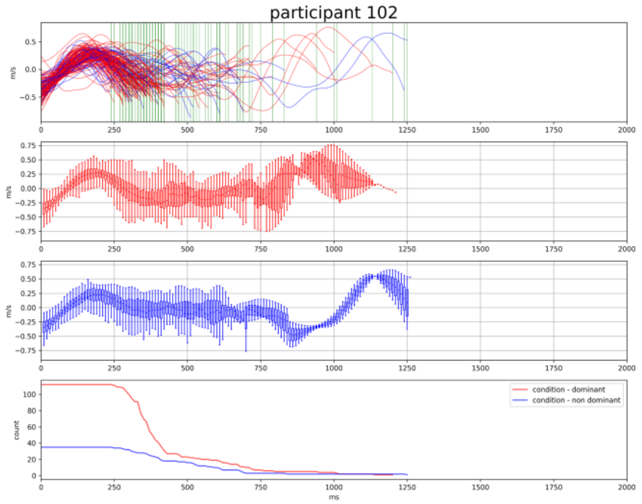
participant 102
m/s
count
ms
condition - dominant
condition - non dominant

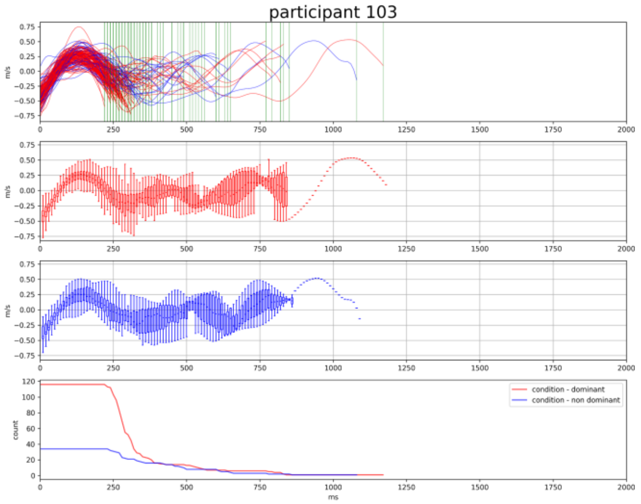
participant 103
m/s
count
ms
condition - dominant
condition - non dominant

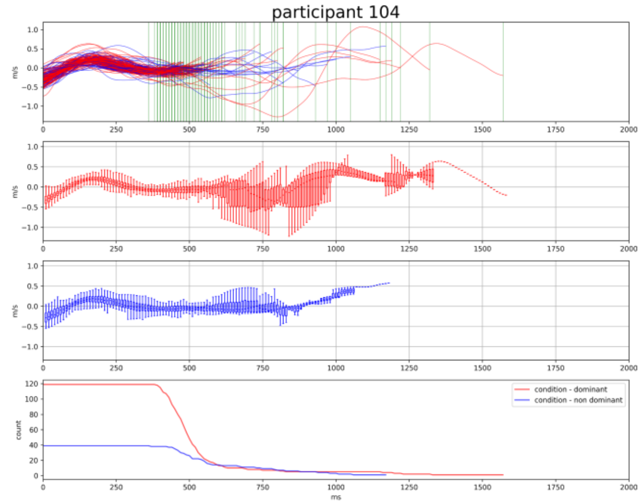
participant 104
m/s
count
ms
condition - dominant
condition - non dominant

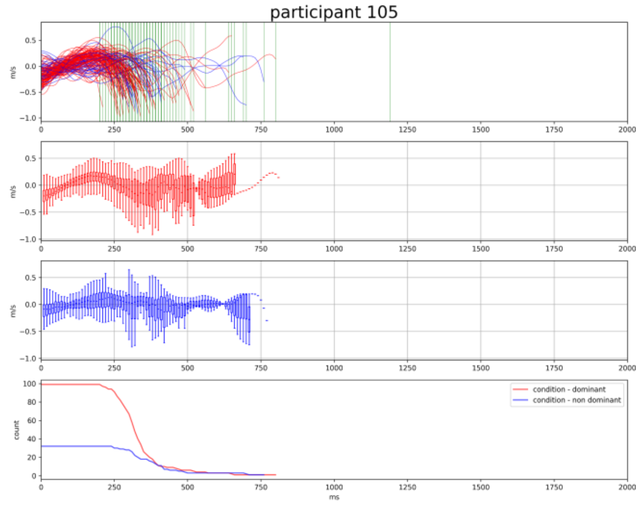
participant 105
m/s
count
ms
condition - dominant
condition - non dominant

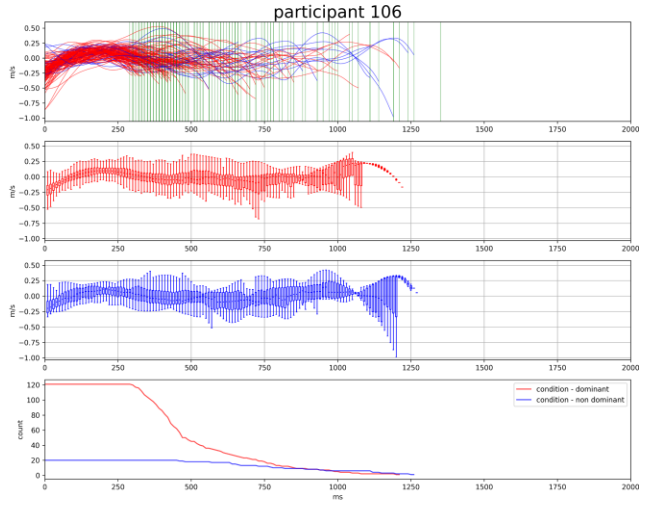
participant 106
m/s
count
ms
condition - dominant
condition - non dominant

participant 107

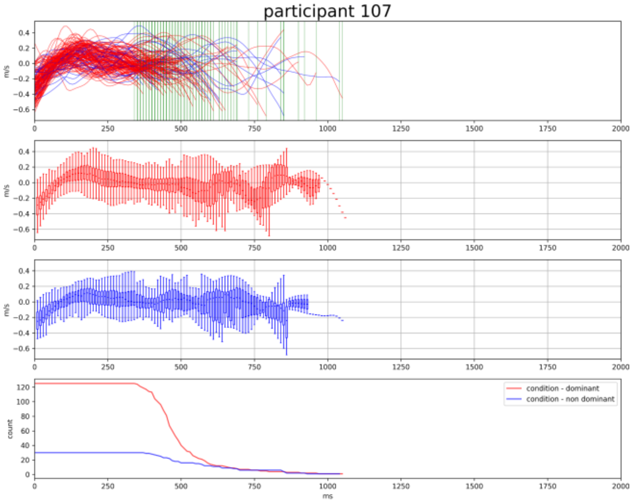


participant 108

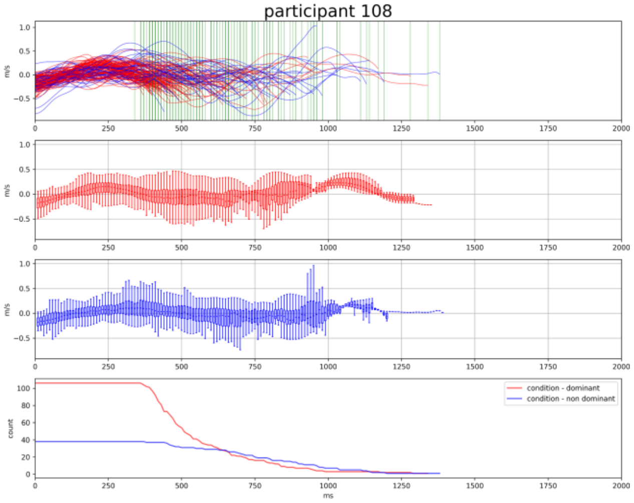


participant 109

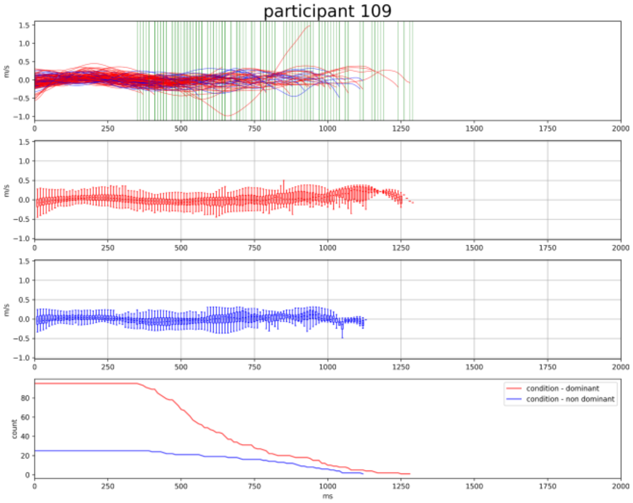


participant 110

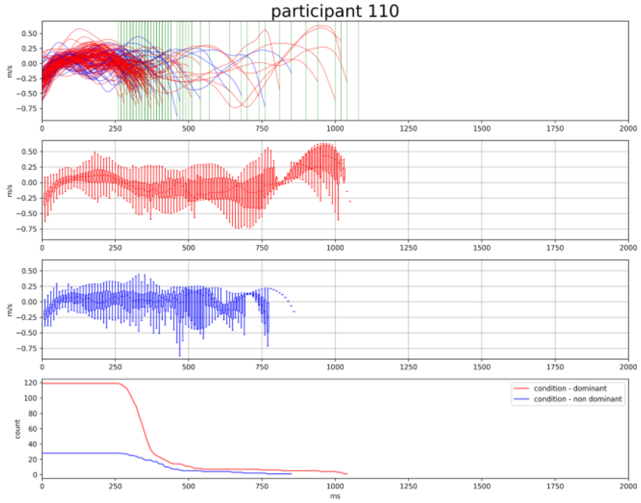


participant 111

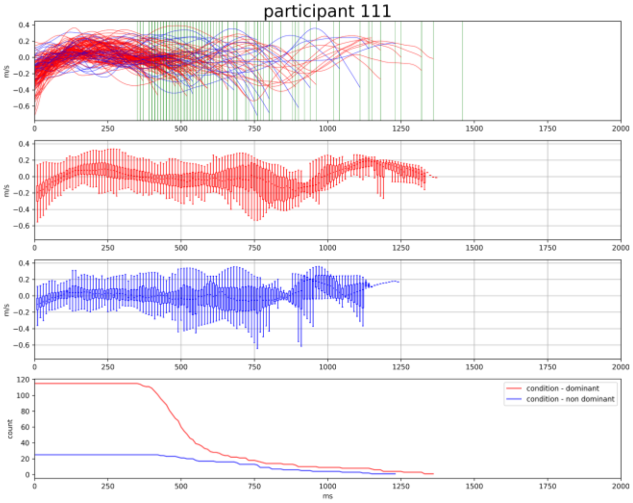


participant 112

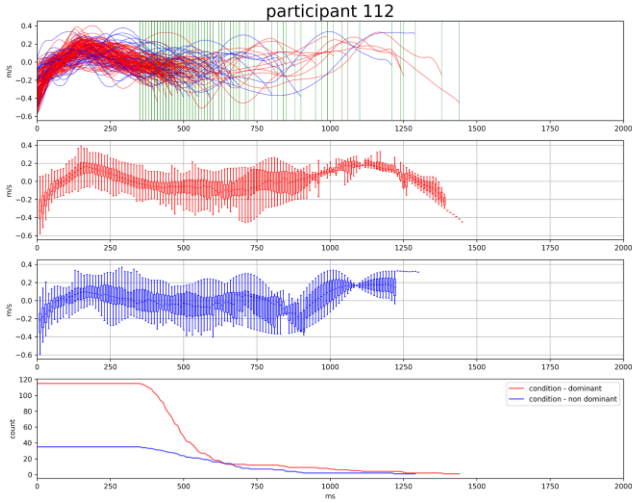

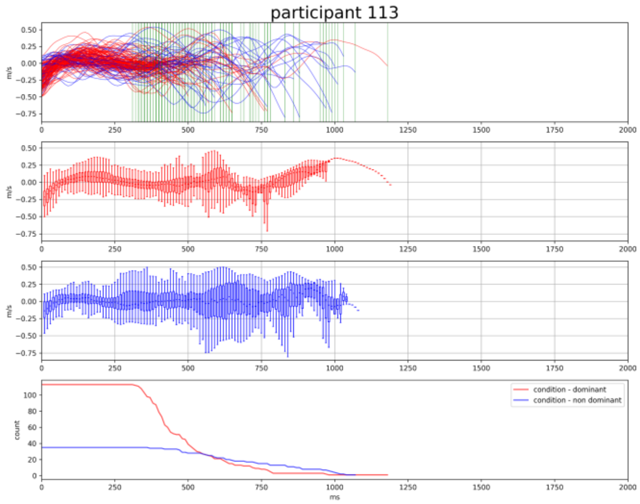
participant 113
condition - dominant
condition - non dominant
m/s
count
ms

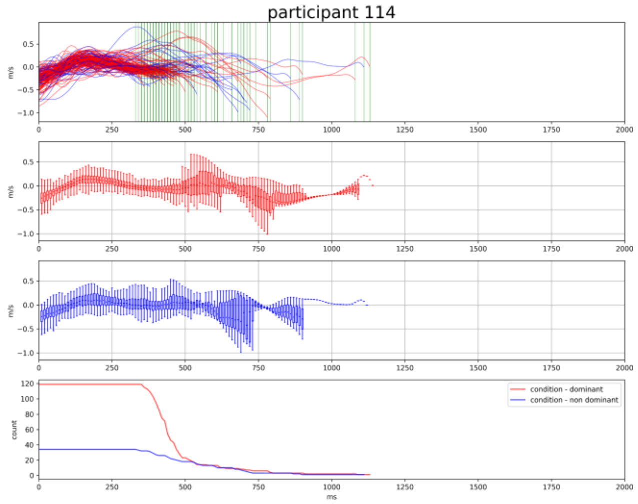
participant 114
condition - dominant
condition - non dominant
m/s
count
ms

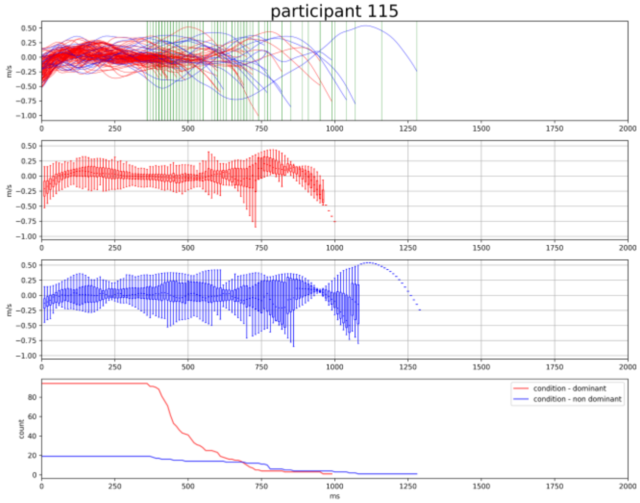
participant 115
condition - dominant
condition - non dominant
m/s
count
ms

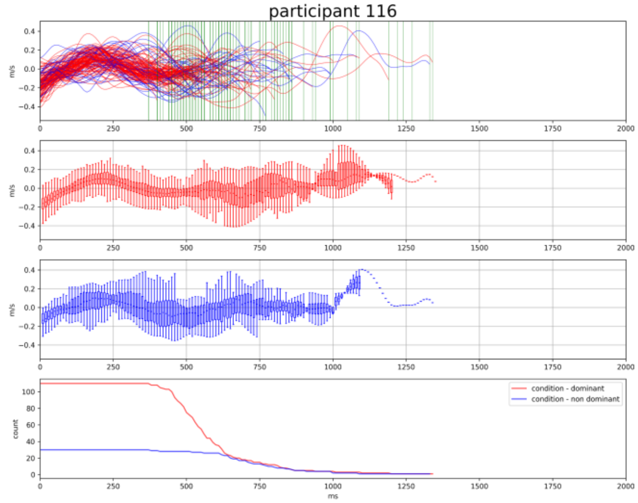
participant 116
condition - dominant
condition - non dominant
m/s
count
ms

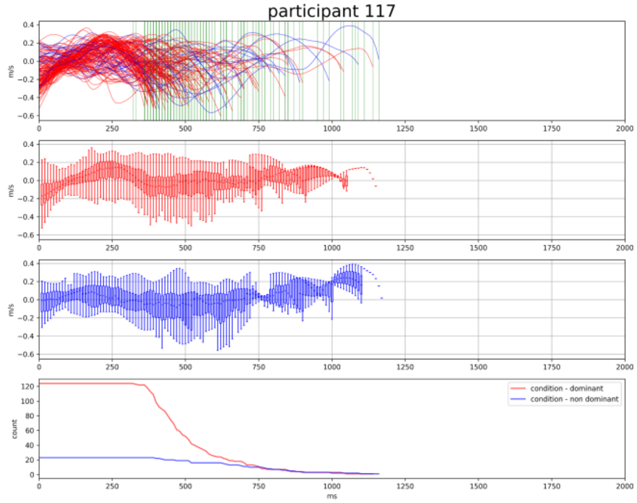
participant 117
condition - dominant
condition - non dominant
m/s
count
ms

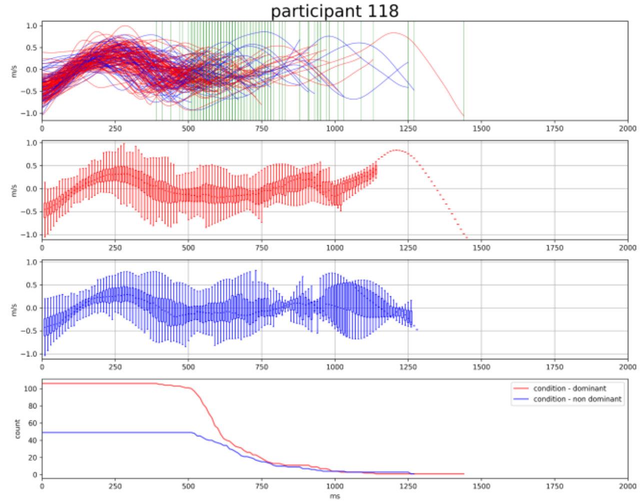
participant 118
condition - dominant
condition - non dominant
m/s
count
ms

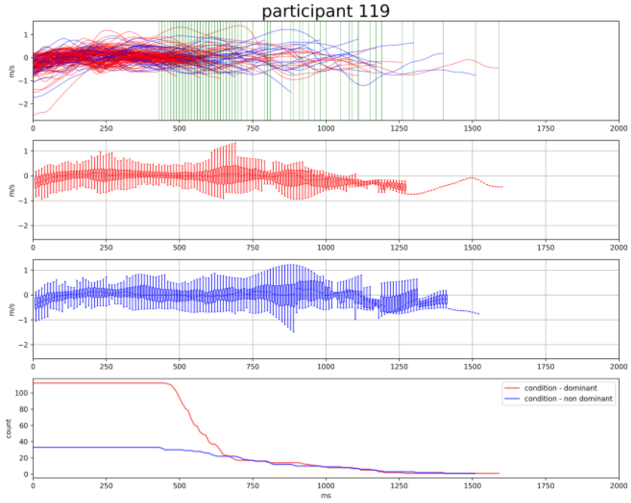
participant 119
m/s
count
ms
condition - dominant
condition - non dominant

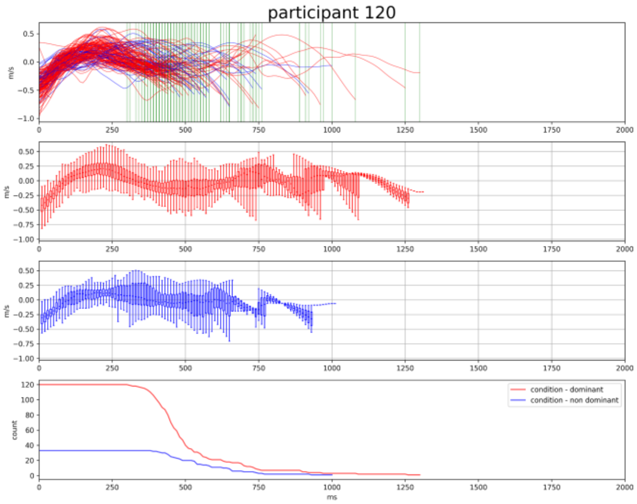
participant 120
m/s
count
ms
condition - dominant
condition - non dominant

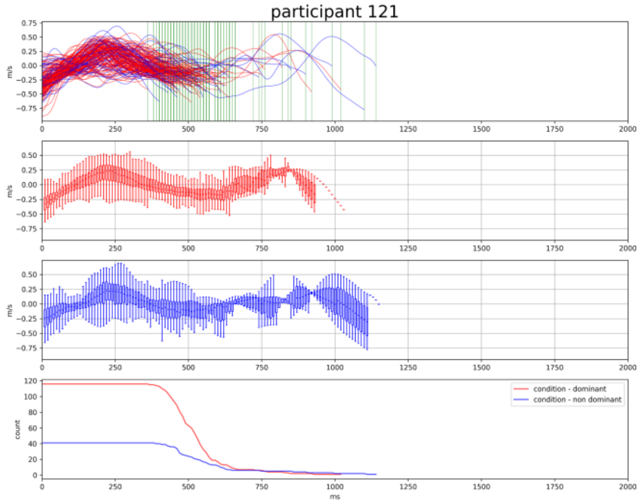
participant 121
m/s
count
ms
condition - dominant
condition - non dominant

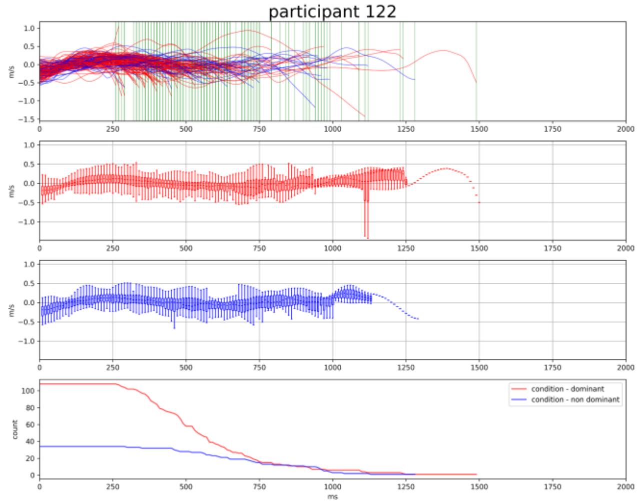
participant 122
m/s
count
ms
condition - dominant
condition - non dominant

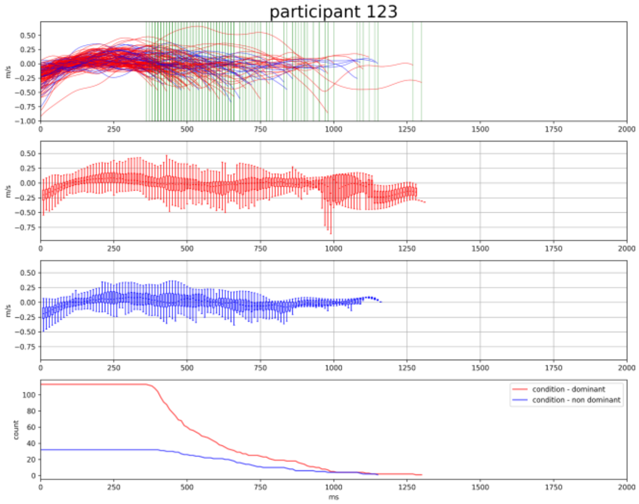
participant 123
m/s
count
ms
condition - dominant
condition - non dominant

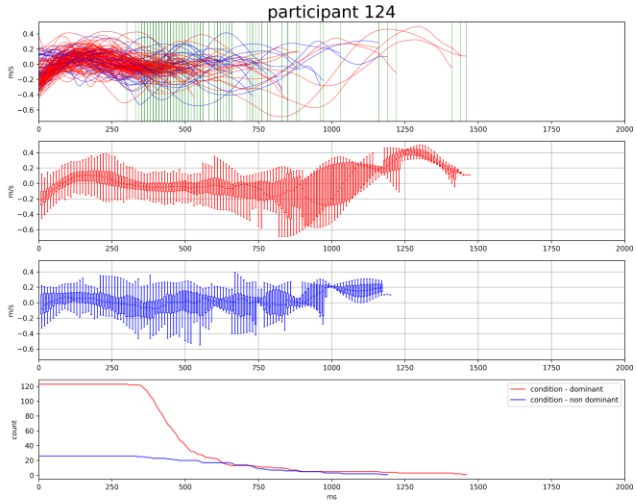
participant 124
m/s
count
ms
condition - dominant
condition - non dominant

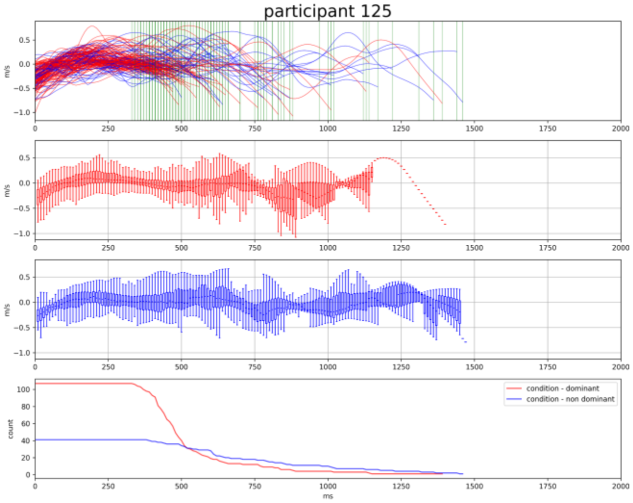


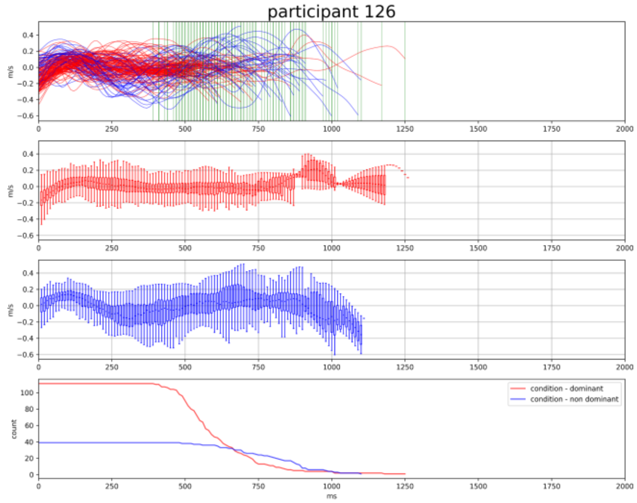


From these graphs we note that, although all participants showed a bell-shaped velocity profile at the beginning of movements, there is a wide intra-group variability. The significance of this motor variability, in relation to the individual characteristics of typically developing children, is largely under-studied in the scientific literature and deserves further investigation to explore the possibility of capturing predictive cues about the children's motor development.

#### 3. ADHD children

Here we report graphical visualisations of individual data from the **ADHD group**. For this cohort of 17 participants, numbers ranging from 1 to 17 are reported in the Figure title as participants' identification code.

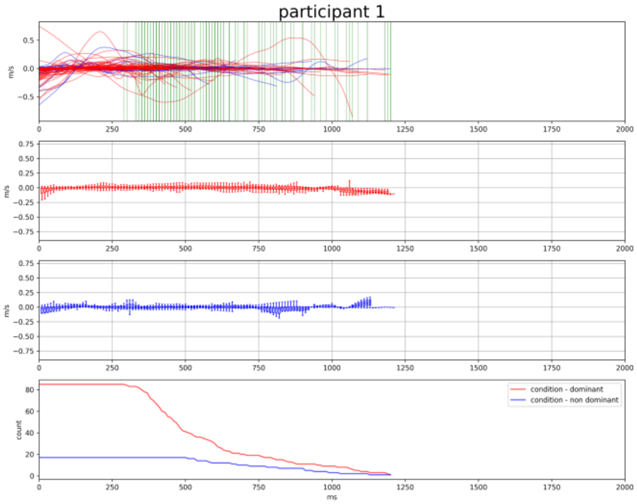


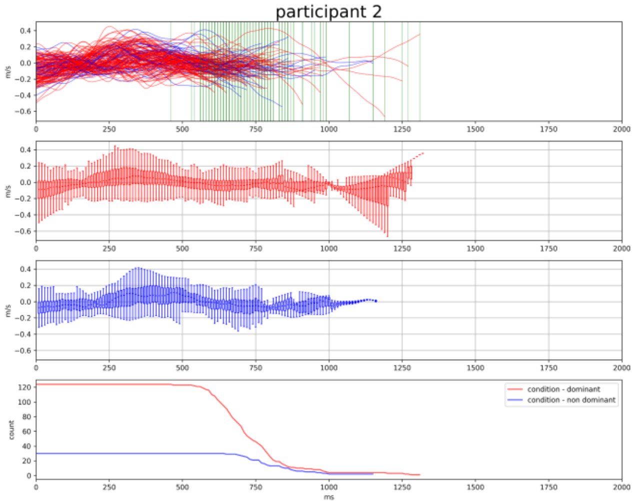

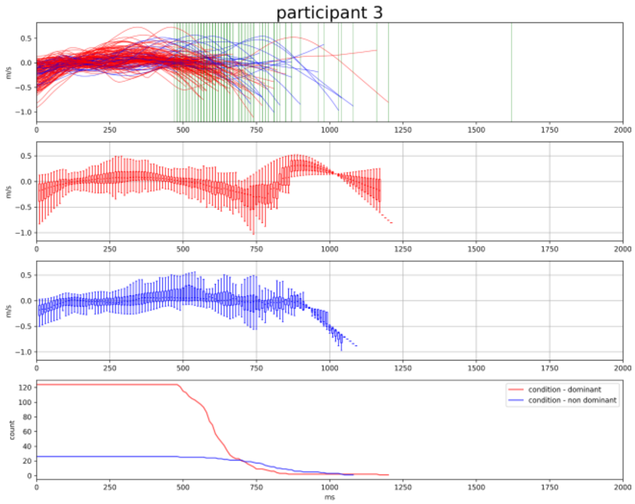
participant 3
m/s
count
ms
condition - dominant
condition - non dominant

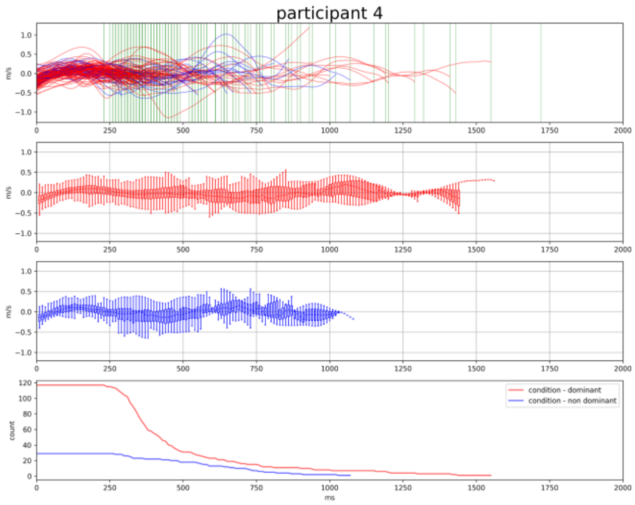
participant 4
m/s
count
ms
condition - dominant
condition - non dominant

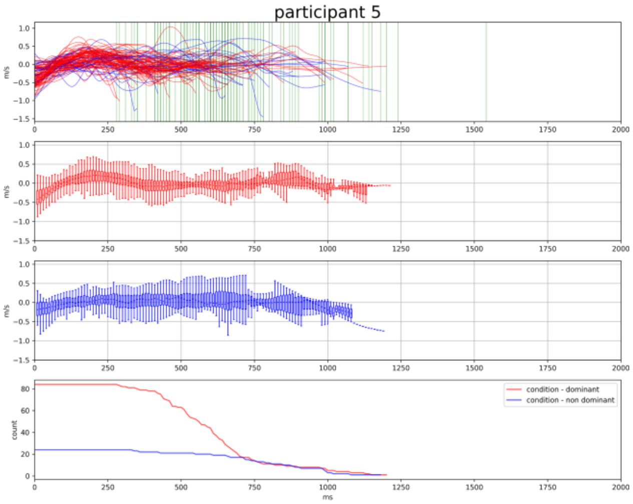
participant 5
m/s
count
ms
condition - dominant
condition - non dominant

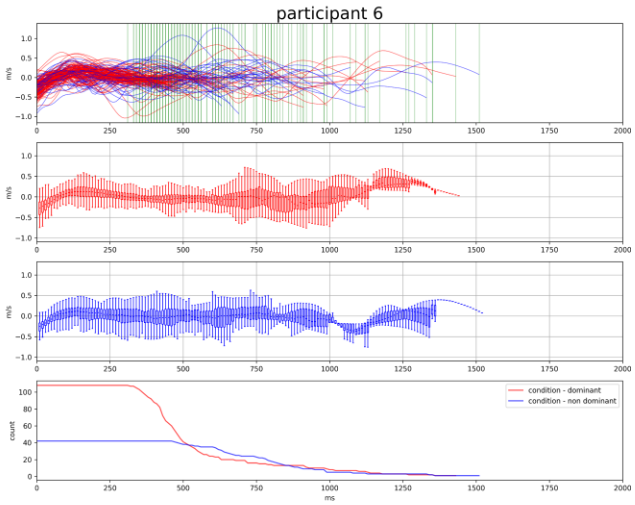
participant 6
m/s
count
ms
condition - dominant
condition - non dominant

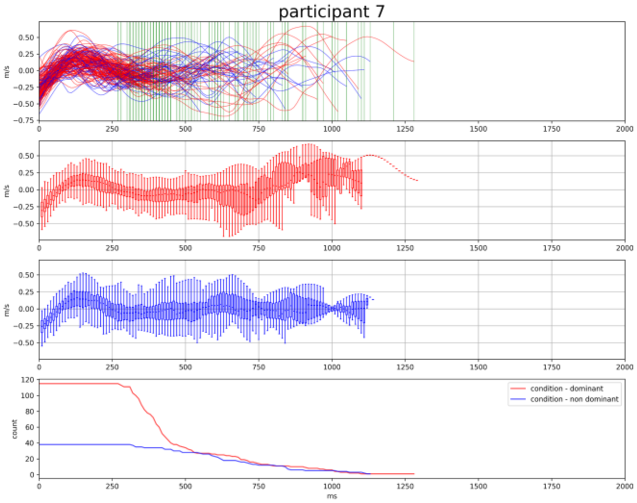
participant 7
m/s
count
ms
condition - dominant
condition - non dominant

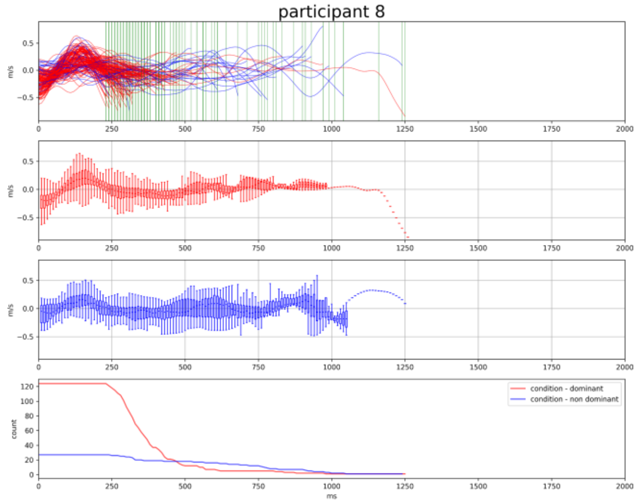
participant 8
m/s
count
ms
condition - dominant
condition - non dominant

participant 9

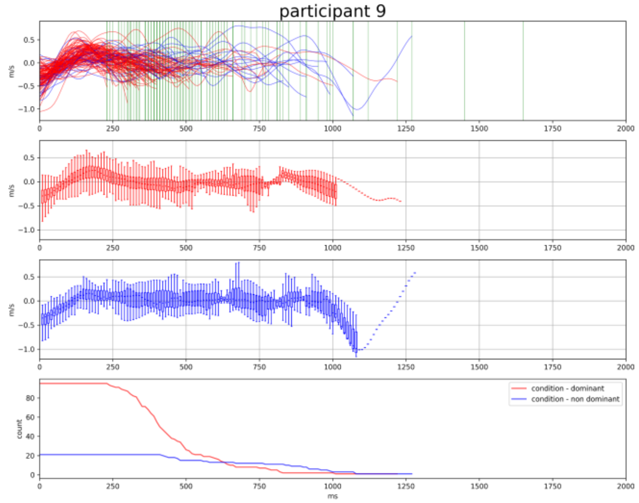


participant 10

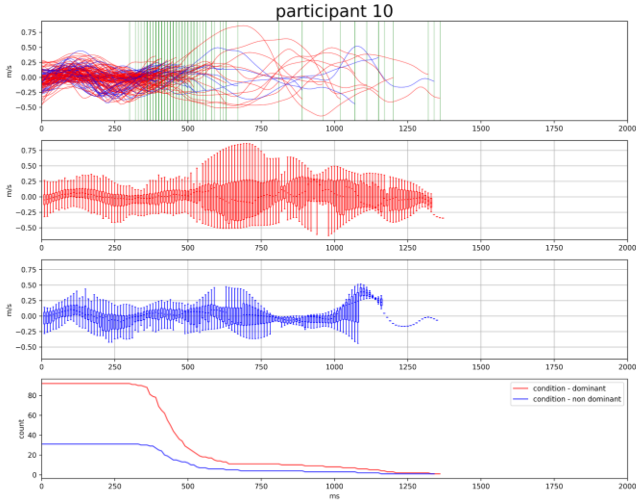


participant 11

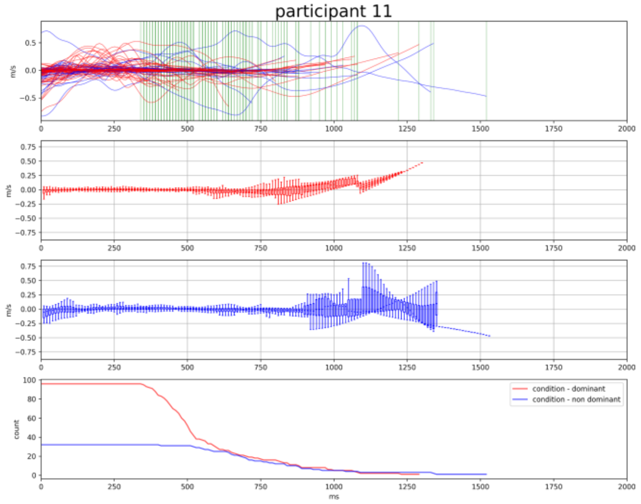


participant 12

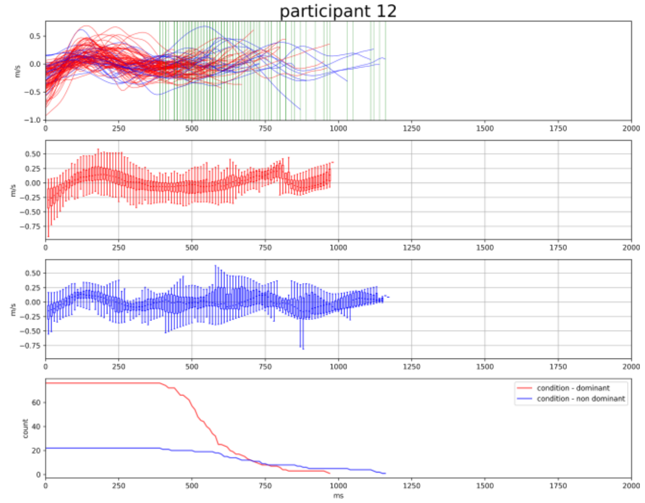


participant 13

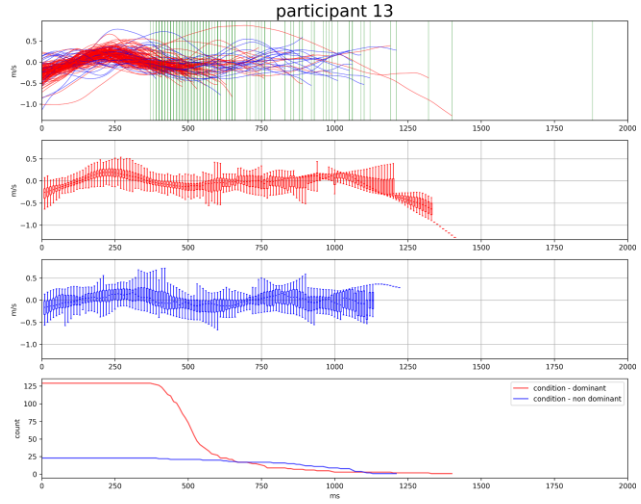


participant 14

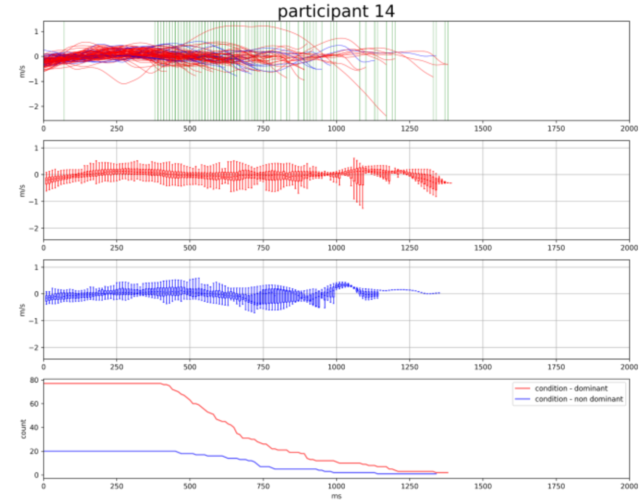

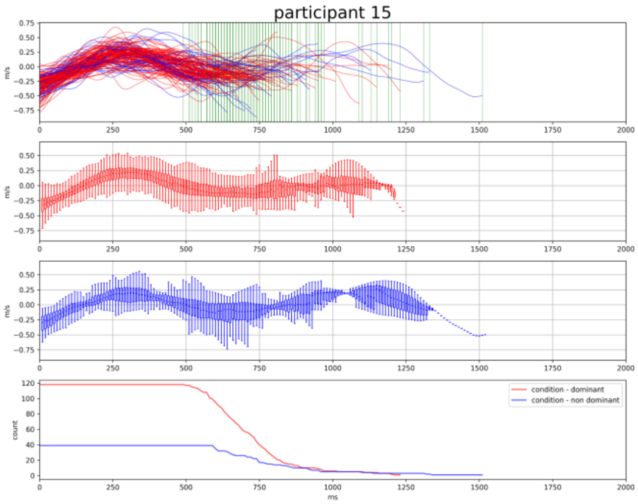


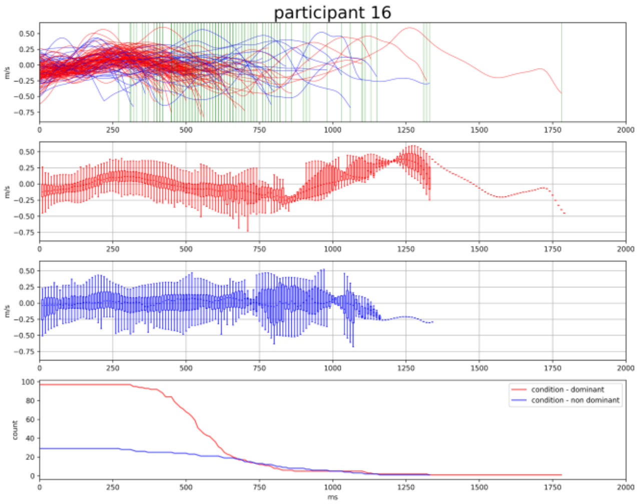


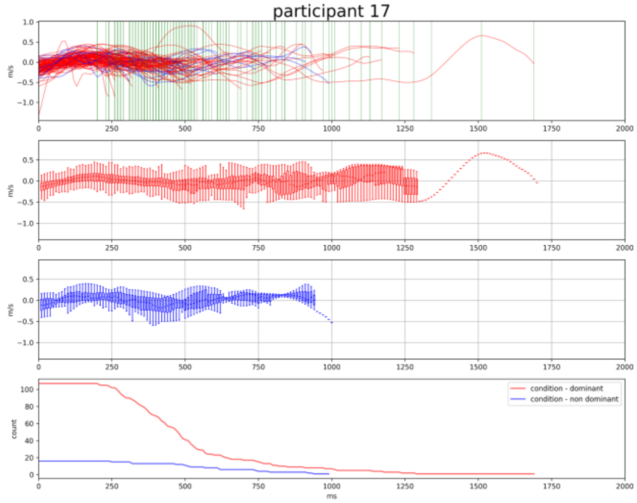


From these graphs we observe that some children from the **ADHD group** did not show an initial bell-shaped velocity pattern (see *participants 1, 2, 3, 11, 14*). As for the TD group, profound intra-group variability is visible and would be worth further investigation. We can speculate that, beyond diagnosis, individual differences in children's motor developmental trajectory interact with other neuropsychological domains to delineate risk profiles that merit clinical attention.
